# Identifying Synergistic Interventions to Address COVID-19 Using a Large Scale Agent-Based Model

**DOI:** 10.1101/2020.12.11.20247825

**Authors:** Junjiang Li, Philippe J. Giabbanelli

## Abstract

There is a range of public health tools and interventions to address the global pandemic of COVID-19. Although it is essential for public health efforts to comprehensively identify *which* interventions have the largest impact on preventing new cases, most of the modeling studies that support such decision-making efforts have only considered a very small set of interventions. In addition, previous studies predominantly considered interventions as independent or examined a single scenario in which every possible intervention was applied. Reality has been more nuanced, as a subset of all possible interventions may be in effect for a given time period, in a given place. In this paper, we use cloud-based simulations and a previously published Agent-Based Model of COVID-19 (`Covasim`) to measure the individual and interacting contribution of interventions on reducing new infections in the US over 6 months. Simulated interventions include face masks, working remotely, stay-at-home orders, testing, contact tracing, and quarantining. Through a factorial design of experiments, we find that mask wearing together with transitioning to remote work/schooling has the largest impact. Having sufficient capacity to immediately and effectively perform contact tracing has a smaller contribution, primarily via interacting effects.

## 1 Introduction

First reported in December 2019 in Wuhan, China [63], the emergence of COVID-19 caught the world by surprise. While scientists were mobilized to understand its pathology, the virus spread to all corners of the world, leading the World Health Organization (WHO) to declare a global pandemic in March 2020 [63]. A year later, the pandemic is still in full swing. COVID-19 is estimated to be the third leading cause of death for the United States in 2020 [12], with a forecasted number of 300,000 deaths attributable to COVID-19 by the end of year [9]. The Centers for Disease Control and Prevention (CDC) estimates that about 53 million infections occurred in the US by the end of September, with only 1 of every 7 being nationally reported [54]. It is thus of paramount importance to identify and implement the *right set* of interventions to effectively slow down the disease progression while waiting for the distribution and intake of a vaccine.

Mathematical models of infectious diseases commonly use ‘compartmental models’, which are systems of coupled differential equations that predict global quantities such as the number of infections at any given time [26, 30, 50, 66]. Agent-based models (ABMs) were later used to capture heterogeneity in populations [64], representing how different individuals (e.g., in age, gender, or socioeconomic factors) have different risks, or willingness and abilities to comply with preventative measures [25]. In the absence of a widely used vaccine, our study and previous ABMs rely on *non-pharmaceutical interventions* including individual-level preventative measures (e.g., stay at home, social distance, using face masks) and subsequent interventions (e.g., contact tracing, quarantining).

As evidence in table 1, there are several limitations to these studies. First, and most importantly, previous modeling efforts only considered the effects of a *small subset of interventions* commonly adopted by national governments, and not all interventions are applied in conjunction. In a sample of 10 studies, we note that an average of only 2.3 interventions are used simultaneously, which is significantly less than the number of interventions that have been implemented or considered by governments [51]. Second, when several interventions are implemented, there is limited analysis to assess whether synergistic effects are obtained or whether most of the benefits can be attributed to only some of the policies. Identifying the right set of synergistic interventions is an important information for policy-making, particularly as compliance becomes an issue. Finally, we frequently note reporting issues such as an insufficiently motivated number of runs (since stochastic models need replicated runs to achieve a sufficient confidence interval) or a coarse resolution when the target population is large.

**Table 1.**
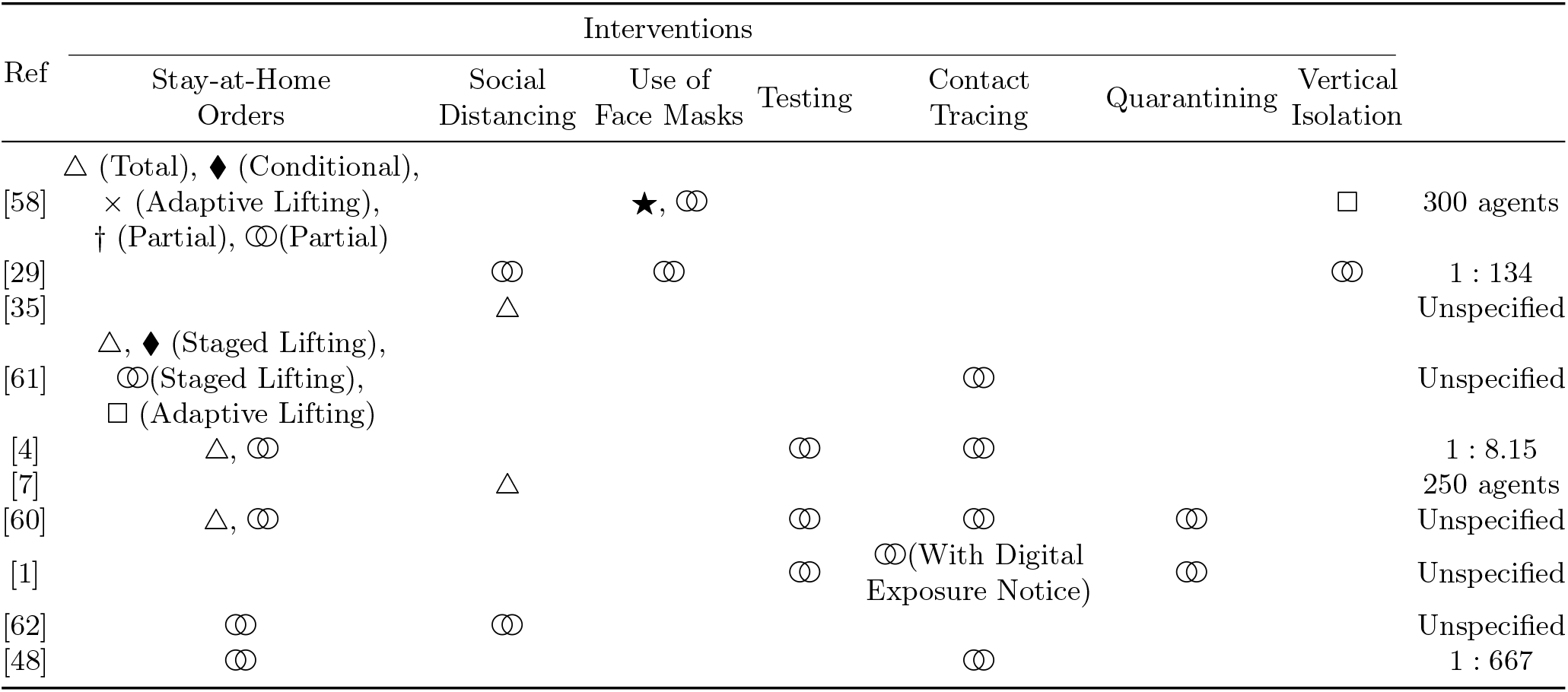
Survey of agent-based COVID-19 models and the interventions they considered. For each row, the number of unique markers gives the number of different scenarios considered in the study. Interventions that were applied in conjunction are denoted with the 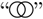 symbol. Each study had only one such scenario. Comments regarding each intervention and scenario are given in parentheses.

To support policymakers in identifying the right set of interventions and provide the necessary confidence in high-stake computations, our paper uses Design of Experiment (DoE) techniques and large-scale cloud-based simulations that measure the individual and interactive effects of interventions at a detailed level. Specifically, our contributions are twofold:

— We measure the impact of *six interventions* on disease prevalence and mortality by accounting for interactive effects. Our interventions include face masks, social distancing, stay-at-home orders, testing, contact tracing, and quarantining. We simulates these interventions at *various levels of adherence*, thus accounting for possible variations in behavioral responses.
— We provide all results at a very accurate population scale of 1 : 500 for the USA and within a confidence interval of at least 95%. This requires a massive number of computationally intensive experiments, which are performed through the cloud via the Microsoft Azure platform.

The remainder of the paper is structured as follows. In section 2, we provide an overview on modeling the biology and interventions regarding COVID-19. In section 3, we describe the procedures of our experiments, including the principles of factorial designs of experiments. Our results are discussed in section 4.

## 2 Background

Once a human host is infected with COVID-19, the disease self-replicates for an amount of time known as the *incubation period*. It is currently estimated that on average, this period lasts for 5.1 days, with a range from 2 to 14 days [38]. Once the viral load has reached a sufficiently high level, symptoms may occur and the host becomes able to spread the infection [10]. Approximately 55% of all infections are considered symptomatic [27] and vary in severity [14]: most are mild and do not require hospitalization for recovery, but severe cases require the support of an Intensive Care Unit (ICU) such as mechanical ventilation [27]. The course of the disease depends on a variety of risk factors such as age, whereby more senior members of society have higher hospitalization rates [11].

During the first phase of COVID-19 (January-June 2020), most COVID-19 models (57%) focused on capturing the transition from a healthy state to being infectious and eventually recovering or dying [22]. This category is known as ‘mass action compartmental models’ because the population is conceptually grouped into categories or ‘compartments’. In the case of COVID-19, the population can be abstracted into compartments such as Susceptible (S), Exposed (E), Infectious (I), or Recovered (R). The model is then described by a system of differential equations stipulating the transitions in and out of the various compartments based on observed rates, such as the contact rate or the rate of exposure per contact. Models developed under this approach are usually characterized by the categories that they capture, listed in the order with which individuals can move from one category to another (e.g., the SEIR from the MIT^1^, `Rainier`, or Columbia^2^). A variant known as ‘structured metapopulation models’ can represent differences in contact patterns by dividing the compartments to reflect sub-populations. For example, sub-populations can be based on age (to reflect the different risk levels and behavior as the age increases) and/or location (to model the spread of a contagion via commuter flows) [2].

Although compartmental models or structured metapopulation models *start* to support the simulation of interventions (e.g., by lowering the contact rate to represent the impact of social distancing), they are still limited in their accuracy as they only represent groups of individuals. In contrast, Agent-Based Models (ABMs) represent each individual explicitly. This fine-grained resolution is motivated by the important individual differences for COVID-19 in terms of risk factors [14, 65] (e.g., respiratory disease, cardiovascular disease) or access to care [8]. Differences are also pronounced when simulating interventions, as various personal beliefs and values result in heterogeneous behaviors [24] regarding actions such as wearing masks or staying at home. An ABM typically serves in public health as a virtual laboratory in which to simulate the expected effect of detailed intervention scenarios, either on disease incidence and mortality, or on the economy [34]. For instance, a COVID-19 ABM was used in April to examine varying quarantine durations and approaches to lifting the quarantine in New York City [28]. Similarly, an ABM was produced to forecast disease incidence and mortality based on various ongoing interventions (e.g., physical distancing) in France [29]. As a growing number of ABMs were developed over the last few months, authors have posited that “ABMs have been utilized in the majority of research” [18]. After a proliferation of independently developed models, we started witnessing the emergence of COVID-19 ABM modeling *packages* or toolkits, such as `COMOKIT` [16] or `Covasim`, developed by the Institute of Disease Modeling [36]. In this paper, we use `Covasim` for two reasons. First, this toolkit is fully transparent by being open source with adequate documentation. This transparency has been emphasized early on [6] and cautionary tales showed its necessity, as high-profile models which were often cited by government agencies turned out to contain bugs and problematic assumptions once the code was later released and inspected [32]. Second, the toolkit has been used in several peer-reviewed studies [48, 57], which provides an additional layer of scrutiny and higher quality expectations than unpublished e-prints (arXiv, medRxiv) or ‘preprints’.

The intervention strategies simulated by an ABM have necessarily been restricted to non-pharmaceutical approaches, in the absence of a vaccine. Such approaches can be further divided as follows:

— **Preventative Care**. Educate the public on practices to reduce the chance of infection. Encourages social distancing, frequent hand washing, and the use of personal protective equipment such as face masks [53].
— **Lockdown**. Reduces the mixing of susceptible and infectious populations. Concrete measures include school closures, workspace closures, and travel restrictions [41].
— **Testing**. Identifies infected individuals (including asymptomatic cases) and places them on quarantine to stop further infections. Contact tracing also often follows from positive tests to further limit the spread of the virus.

One of the key advantages of using `Covasim` to accurately forecast the effects of the interventions above is the handling of *networks*. As explained with metapopulation models, there are different frequencies and duration of interactions between people of different ages or living in different locations. In addition, the probability of passing on the virus depends on the intimacy of the interactions, such that an asymptomatic infected person is more likely to spread the infection to family members than to co-workers. This results in ‘substantial transmission heterogeneities’, as evidenced by detailed contact tracing data [59]. To accurately model policies such as contacting tracing [52] or balancing social groups [45], it is thus essential to simulate the interactions of virtual agents over synthetic networks that share characteristics with the structure of real-world social networks [15]. `Covasim` accounts for these effects by placing agents in four networks (work, school, community, home), each representing a certain type of contact with different levels of infectivity. Using these networks, we can simulate a ‘working-from-home’ policy by reducing the number of edges in the work network, or school closures by eliminating the school network.

## 3 Methods

### 3.1 Using the Covasim platform

Our simulations are performed using the `Covasim` platform as explained in our background section. We take two specific actions to ensure the accuracy of the simulations within our application context. First, we use a resolution of 1 : 500 (i.e. each simulated agent represents 500 real-world people) which, given the US population of 328 million people, gives a simulated agent population of over 650, 000. This positions our paper among large-scale COVID-19 simulations, which exceed half a million agents [3]. Second, we used the Confidence Interval method [55, pp. 184–186] to identify and perform a sufficient number of replications in each scenario such that the average results are within a 95% Confidence Interval (95% CI). For additional examples of the CI method, see [21, 23].

Our goal is to identify the most effective set of interventions that can control the spread of COVID-19 in the United States. To that end, our simulation scenarios correspond to combinations of interventions at various levels (detailed in subsection 3.2). To quantify the effects of stipulated interventions, we report the total infections after 180 days. It is important to note that our simulated time period starts in September 2020 rather than at the beginning of the pandemic, when most of the population was susceptible and almost none had recovered. By choosing a more current starting time, our simulated dynamics take into account the presence of recovered agents as well as individuals who are already at various stages of the infection. We made two modifications to `Covasim` accordingly: randomly selected agents are set to recovered based on the amount estimated by the CDC and scaled to our population resolution; and a separate random subset of agents are set to infected, with their date of infection chosen among the 14 days preceding the start of the simulation (also in accordance with CDC data) such that agents are realistically set to different stages of infection.

As `Covasim` already contains a large number of population parameters calibrated for the US, we used the same default values in our work. There were three exceptions, as the knowledge base upon which `Covasim` was built has since evolved and needed to reflect our current understanding of real-life disease dynamics. The following three modifications were implemented as follows:

1. Based on recent evidence, the distribution of incubation period is such that the mean is at 5.1 days, the range is from 2 to 14 days, and 97.5% of people develop symptoms within 11.5 days [10,38]. In `Covasim`, incubation is broken into two pieces: one from infection to viral shedding (denoted by *s*), and the other from viral shedding to developing symptoms (denoted by *i*). The distributions assumed by *s* and *i* are *lognormal*(4.6, 4.8) and *lognormal*(1, 1), respectively^3^. Upon evaluation, however, the combined distribution does not match the latest evidence on incubation period. Consequently, the distributions of *s* and *i* were adjusted to *lognormal*(4.1, 4.8) and *lognormal*(1, 1.8), respectively. The largest change is the increased standard deviation of symptom onset, which is justified by [10]. This adjustment results in a combined distribution that agrees with the observed values (Figure 1).
2. The literature reports a proportion of symptomatic cases of 0.6 to 0.65 [47], which differs from `Covasim`’s default of 0.7 [36]. Consequently, we scaled down by 6*/*7 to bring the parameters in line with expectation.
3. Testing delays vary based on regions, availability of testing resources and the need for them. However, `Covasim` uses a fixed testing delay that would apply to the whole population. We thus changed it to a distribution based on a recent survey [40] showing that mean waiting times nationally were 4.1 days, with extreme wait times of up to 14 days. We use the same discrete distribution as shown in the report (c.f. Figure 1 in [40]) to reflect the real-world testing delay.

### 3.2 Interventions

Many interventions can be specified in `Covasim`. We focused on four *categories* of interventions: mask wearing (realized as direct reductions of the susceptibility of simulated agents), lockdowns, testing, and contact tracing. Several parameters are required for each category in order to precisely characterize how the intervention will unfold. For instance, testing is a matter of *how many* tests are available on a daily basis, *when* to test individuals who’re entering quarantine as they were exposed to the virus, and the extent to which tests are *reliable* (i.e. test sensitivity). We list the parameters for each category of intervention in Table 2 together with the range of values that *could* be used and the specific subset that we do use. Our choice is motivated by the references provided in the table and is often limited to a binary due to the experimental set-up explained in the next subsection. Every intervention is applied for the entire duration of the simulation.

**Table 2.** `Covasim` interventions used in this paper. Interventions parameters, values, and sources are provided. The following shorthands for networks are used: *w →* work network, *s →* school network, *h →* home network, and *c →* community network.

| Intervention | Parameters | Possible Values | Values Chosen | Var. | Ref. |
| --- | --- | --- | --- | --- | --- |
| Mask Wearing | Fractional reduction in transmissibility | $[0, 1]$ | 80% | | [13] |
| | Applied networks | $\mathcal{P}(\{w, s, h, c\})$ | $\{\{w, s, h, c\}, \{w, s\}, \{c\}\}$ | $X_1$ | |
| Lockdowns | Fractional reduction in contacts for $\{w, s\}$ | $[0, 1]$ | $\{5\%, 30\%\}$ | $X_2$ | [31] |
| | Fractional reduction in contacts for $c$ | $[0, 1]$ | $\{10\%, 30\%\}$ | $X_3$ | [31] |
| | Applied networks | $\mathcal{P}(\{w, s, h, c\})$ | $X_1 \setminus h$ | | |
| Testing | Number of tests per day | Any integer | $\{6, 11.1\} \times 10^5$ | $X_4$ | [33] |
| | When to test quarantined individuals | $\{\text{start, end, both, daily}\}$ | $\{\text{start, end, both}\}$ | $X_5$ | |
| | Test sensitivity | $[0, 1]$ | $\{55\%, 100\%\}$ | $X_6$ | [42] |
| Contact Tracing | % of contacts of a tested person that can be traced | $[0, 1]$ | $\{0.2, 1\}$ | $X_7$ | [37] |
| | Delay in contact tracing | Any integer | $\{0, 7\}$ | $X_8$ | [37] |
| | Start contract tracing without waiting for results | $\{\text{True, False}\}$ | $\{\text{True, False}\}$ | $X_9$ | |

**Fig. 1.**
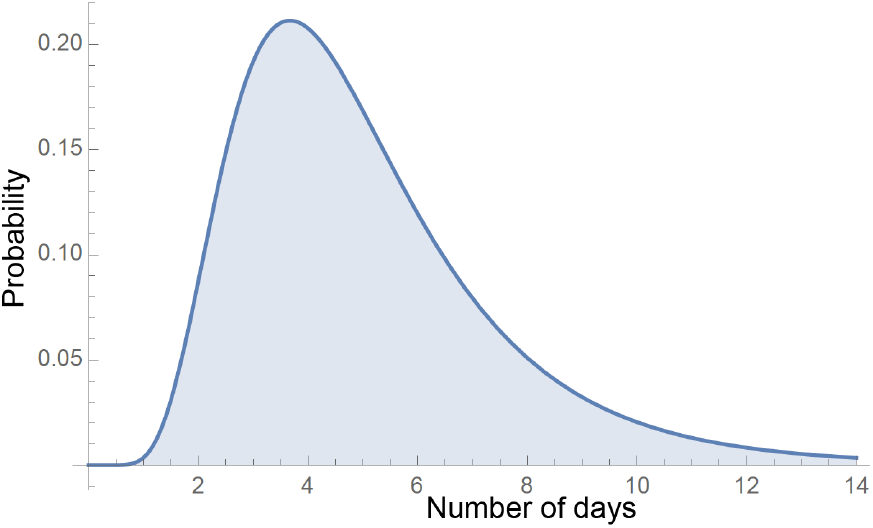
Probability distribution function (PDF) of incubation period with adjusted parameters to reflect recent evidence [10, 38].

### 3.3 Factorial Design

Our goal is to measure the impact of interventions on disease prevalence. An *ineffective* Design of Experiments (DoE) would be to implement only one intervention at a time, which would overlook the *interactions* among interventions that may contribute to making a *set* stronger than the sum of its parts. However, it is computationally infeasible to try all combinations of values. For example, several interventions are specified by continuous values in the range [0, 1]: the reduction in transmissibility via mask wearing, the reduction in contacts, or the percentage of contacts that can be tested. Even if only ten samples were used for each range, the number of combinations is multiplicative (10 *×* 10 *×* 10 = 1, 000) hence the experimental space grows exponentially in the number of parameters (‘full factorial design’). We thus use a common intermediate known as a 2^*k*^ factorial design in which each parameter is set to two values, designated as ‘low’ and ‘high’ [44, pp. 233–303]. For an overview of DoE for Agent-Based Models, including gridded designs such as the 2^*k*^ factorial, we refer the reader to the work of Susan M. Sanchez [56]. For detailed applications, see [19, 20].

Our four categories of interventions result in 9 parameters (Figure 2), which are listed as *X*_1_, …, *X*_9_ in Table 2 together with their low and high values. A factorial design serves to investigate the synergistic effects of these parameters, by measuring the response *y* (average number of infections at the end of the simulation) for each simulated combination. As mentioned in section 3.1, *y* is obtained over a number of replications necessary to fit the 95% confidence interval to within 5% of the average. To determine the contribution of each parameters (individually as well as in groups), we compute the variances in response contributed by each combination and perform an *F* -test. In other words, the variance is decomposed over individual parameters (*X*_1_, …, *X*_9_) as well as interactions of parameters. We speak of 2nd order interaction when examining the joint effect of two parameters (*X*_1_*X*_2_, *X*_1_*X*_3_, …, *X*_8_*X*_9_), 3rd order interaction for three parameters, and so on. We include up to 3rd order interactions to scan for possible effects that would only happen when three interventions are applied jointly. In line with our previous studies, higher level interactions are not considered as the cost of computing and storing them is not justified by their diminishing effects [39].

**Fig. 2.**
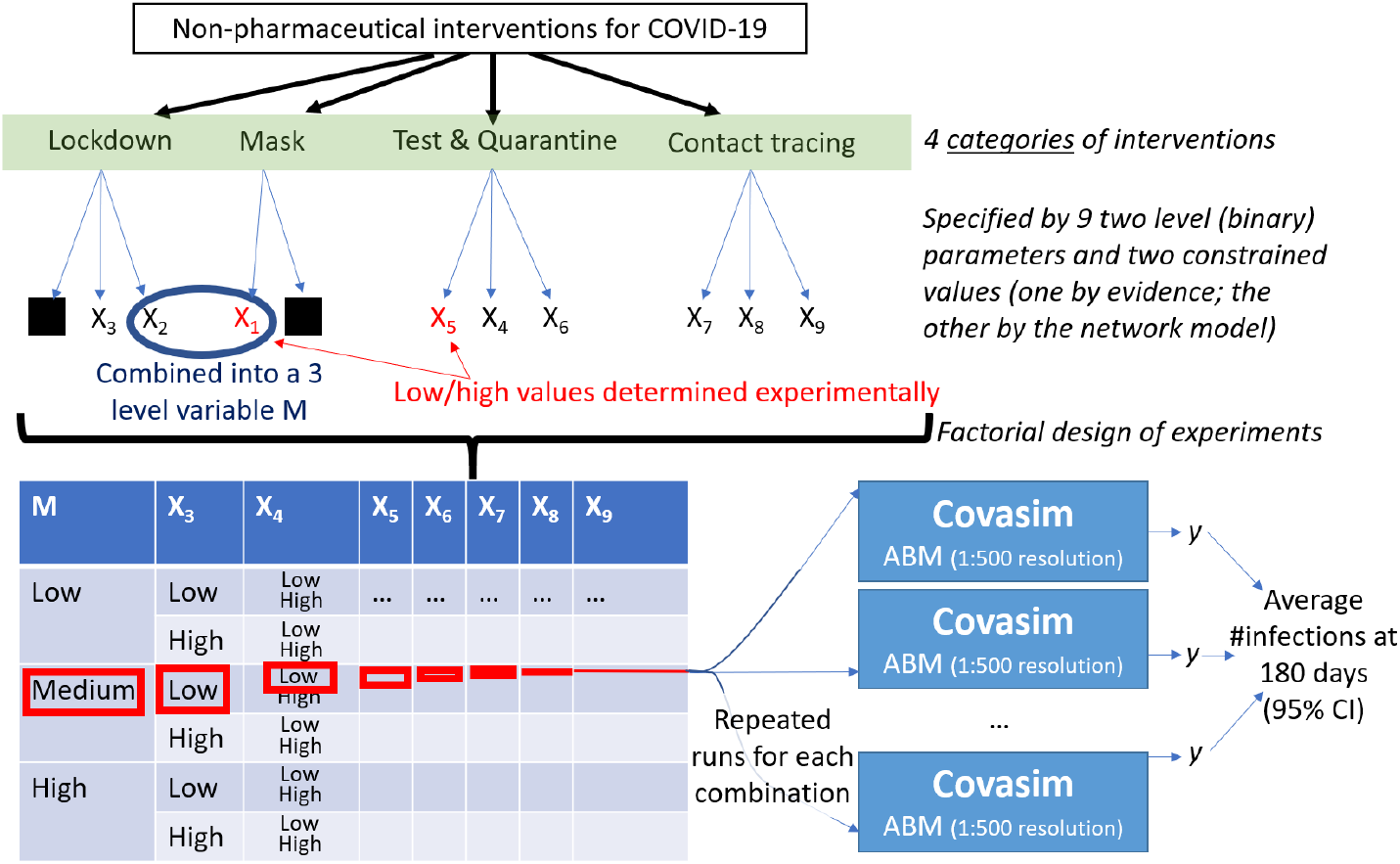
Process flowchart from the identification of interventions (top) and their operationalization in the simulation to the factorial Design of Experiments (DoE) and repeated runs of a stochastic model targeting a 95% Confidence Interval.

There are two special cases when applying the 2^*k*^ factorial design to COVID-19 interventions. First, we cannot assume which values of *X*_1_ (networks for mask wearing) and *X*_5_ (whether to test at the start, end, or both times in a quarantine) have the highest or lowest impact on disease incidence. Consequently, we performed simulations for each of the three possible values of *X*_1_ and *X*_5_. We identified the maximum and minimum number of infected individuals across these simulations, and thus set the high and low values accordingly (Table 3). For example, consider that we have 10 sick individuals when testing at the start of quarantine, 100 when testing at the end, and 1000 when testing at both times. In this case, ‘start’ is the intervention with highest impact 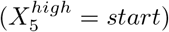 while ‘both’ does the least to control the spread of the virus 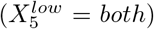. Second, not all 2^9^ = 512 combinations of the high and low levels are valid because of the interdependency between the networks for mask wearing and lockdown. If an intervention applies to work and school (e.g., working and learning remotely) then the fractional reduction of contact in the *community* should be 0 rather than 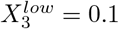, since the community network is not concerned by the intervention.

**Table 3.** Maximum responses caused by each choice of variables *X*_1_ and *X*_5_.

| Variable | Value | Max Response | Level |
| --- | --- | --- | --- |
| $X_1$ | $\{w, s, c, h\}$ | 678.93 | High |
| | $\{w, s\}$ | 278571.55 | — |
| | $c$ | 359937.49 | Low |
| $X_5$ | start | 359862.78 | — |
|  | end | 359606.72 | High |
|  | both | 359937.49 | Low |

To resolve this issue, we first note that the highest impact intervention is the one that targets all networks, hence 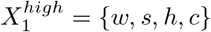 as seen in Table 3. The core question is thus the *low* value, which is either {*w, s*} or *c*. This choices sets either *X*_2_ or *X*_3_ (resp. the fractional reduction in contacts for *c* and {*w, s*}) to 0%. Consequently, 2^7^ = 128 combinations are invalid and 384 = 3 *×* 2^7^ remain. To avoid the complications of an incomplete parameter space, we combine two parameters into a new variable *M* with 3 levels (Table 4) and keep the rest unchanged. We can interpret this variable to be the level of “social distancing” in the broadest sense that is enforced. Moreover, the problem reduces to an ordinary factorial design analysis with one 3-level factor, which is analyzed using established methods [44, pp. 412–414].

**Table 4.**
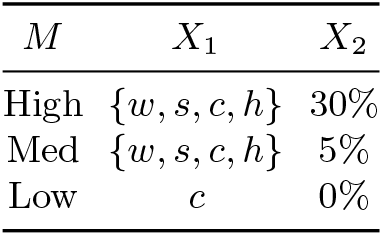
Correspondence between levels of *M* and combinations of levels of *X*_1_ and *X*_2_, given *c* as the low level for *X*_1_.

| $M$ | $X_1$ | $X_2$ |
| --- | --- | --- |
| High | $\{w, s, c, h\}$ | 30% |
| Med | $\{w, s, c, h\}$ | 5% |
| Low | $c$ | 0% |

## 4 Results and Discussion

Our research objective is to measure the individual and interactive effects of COVID-19 interventions through a detailed Agent-Based Model reusing open source code and an approach validated by another research group in several peer-reviewed publications. To provide detailed results (resolution of 1:500, 95% Confidence Interval) while managing the computational load, we conducted our experiments on the Microsoft Azure Platform. Our experimental approach is shown in Figure 2 and analyzed as a factorial Design of Experiments. Table 5 summarizes the most potent combinations of intervention parameters that impact the number of new cases, as well as their actual contributions.

**Table 5.** Factors responsible for at least 1% of the variance in the number of new infection cases

| Variable(s) | Meaning | Contribution (%) |
| --- | --- | --- |
| $M$ | Masks + remote work/school | 66.638 |
| $M$ and $X_7$ | Masks + remote work/school, and ability to trace contacts of tested individuals | 13.759 |
| $X_7$ | Ability to trace contacts of tested individuals | 8.953 |
| $M$ and $X_9$ | Masks + remote work/school, and whether contact tracing starts without waiting for test results | 2.804 |
| $X_9$ | Whether contact tracing starts without waiting for test results | 2.033 |
| $M$ and $X_8$ | Masks + remote work/school, and delay in contact tracing | 1.077 |

Mask wearing together with transitioning to remote work/schooling has the largest impact. It also interacts with the constructs having the next largest impacts, which are the ability to perform contact tracing and whether to start contact tracing without waiting for test results. Our finding about the importance of masks and remote work/school differs from early works (in the first half of 2020) regarding COVID-19. Early systematic reviews released as preprints considered that “the evidence is not sufficiently strong to support widespread use of facemasks as a protective measure against COVID19” and that “masks alone have no significant effect in interrupting spread of [influenza-like illnesses …] or influenza” (cited in [43]). More recently, a systematic review from December 2020 concluded that only four out of seventeen studies ‘supported use of face masks [and] a meta-analysis of all 17 studies found no association between face mask intervention and respiratory infections” [46]. *However, once results are adjusted* for factors such as age and sex, the meta-analysis “suggests protective effect of the face mask intervention” [46]. Similarly, the most recent analyses and commentaries agree that reducing social network interactions in settings such as universities [17] or schools [5] is needed to avoid large outbreaks. Consequently, our result regarding the large effect of masks and remote work/schooling contributes to the more recent evidence base on interventions regarding COVID-19.

Although the preventative approach of using masks and shifting into remote work/school plays the largest role in reducing the likelihood of transmission (by lowering both the number of contacts and the virus transmissibility per contact), we do observe interacting effects with other intervention parameters. In particular, contact tracing is important to mitigate the pandemic, as demonstrated by the case of South Korea [49]. Our study contributes to understanding the specific parameters underlying contact tracing, as results stress the merits of having sufficient capacity to immediately and effectively perform contact tracing.

There are several limitations to this modeling study. First, we are unable to report the contribution of the error factor in the factorial design, that is, the margin due to stochasticity in the model. Evaluating it would require a comparable number of samples for each combination of parameter values, but this number differed greatly among experiments as the 95% Confidence Interval required many more runs for some combinations than others. Running all experiments with the highest number of replications was computational unfeasible, and the wide differences in number of runs precluded a reliable up- or down-sampling. Second, as any model is a simplification of reality, some factors are not represented in the underlying COVID-19 ABM used for this study. For instance, the underlying model does not take into account the duration of immunity, which may be in the scale of a few months depending on initial illness severity [14]. We assumed that immunity would hold during the 6 months (180 days) of simulated time. Finally, although our parameter values were obtained from peer-reviewed sources, the evidence base on COVID-19 continues to evolve and the situation presents new logistical challenges. Some clinical and epidemiological values may thus be refined as additional meta-analyses become available. The logistics of contact tracing at the unprecedented scale of over 200, 000 daily cases may prevent the identification of all contacts, thus lowering what was used as a practically feasible value at the time of this study.

## Data Availability

Our work is based on the Covasim model, which is open source and can be accessed without registration. Table 2 lists all parameters used in this study and tracks their provenance.

## Footnotes

1 https://www.covidanalytics.io/projections

2 https://columbia.maps.arcgis.com/apps/webappviewer/index.html?id=ade6ba85450c4325a12a5b9c09ba796c

3 Here, the *µ* and *σ* denote those of the underlying normal distribution.

